# The effect of smallpox vaccination: unique insights gained from Danish post-honeymoon epidemics 200 years ago

**DOI:** 10.64898/2026.11.16.26368180

**Authors:** Andreas Eilersen, Søren Poder, Marlies Jilles Francine Goedknegt, Jon Bang Ploug, Anders Gorm Pedersen, Romola Davenport, Lone Simonsen

## Abstract

**Background:** Historically, the Variola virus caused deadly smallpox epidemics in children. In Copenhagen, transmission was halted by 1810 when ∼90% of children were vaccinated following the introduction of a mandatory single-dose vaccine program. A 14-year honeymoon period followed, but in 1824 smallpox re-emerged as a milder disease of adults. We set out to study smallpox epidemics between 1824 and 1835 when all smallpox patients in Copenhagen were quarantined.

**Method:** We analyze data from 3795 smallpox patients reported by the treating physicians between 1824 and 1835. The data include age, vaccination status, and outcome. We estimate age-specific case fatality ratios (CFR) and vaccine effectiveness (VE) against mortality.

**Findings:** For the four outbreaks while quarantine was mandatory (1824-1834), the CFR was ∼1% among the vaccinated and ∼25% among the unvaccinated, corresponding to a VE of ∼95%. For the fifth outbreak in 1835, we found that CFR was low among the vaccinated but increased with age, which is evidence of waning vaccine protection. Almost no cases were over 40 years of age, suggesting lifelong immunity from smallpox infection.

**Interpretation:** As the unvaccinated CFR was high and comparable to the pre-vaccine era, the milder post-honeymoon smallpox disease cannot be explained by genetic changes in the virus. Rather, high coverage with an effective vaccine combined with waning vaccine protection in adulthood explains the changing face of smallpox. We demonstrates how high vaccine coverage can disguise an outbreak of highly pathogenic smallpox as the milder Variola minor.

## INTRODUCTION

In the 18^th^ century, smallpox was a dreaded epidemic childhood disease all over the world, and Copenhagen was no exception. The disease killed 10-15% of all young children in the city which at the time was a growing population center ^1,2^, with 87,000 inhabitants around 1770 increasing to just over 100,000 by 1800. The dense population of the city made it especially vulnerable to deadly smallpox outbreaks. The enormous death toll and maiming pock marks pushed people to adopt the practice of inoculation during the 1700s. This was a risky practice in which a small amount of matter from human smallpox pustules was in-oculated into the superficial layers of the skin to induce a hopefully mild illness and subsequent immunity.

Then, in 1796 Jenner developed a cowpox-based vaccine that did not carry these risks. The Danish Professor Winsløw inoculated 80 children in 1801 and saw no serious adverse events. He then challenged two of the children with smallpox and both survived.^3^ A royal vaccination committee was established in October that year and mass vaccination in Denmark began. During 1801-1810 a continually increasing proportion of the birth cohort was vaccinated.^4^ The vaccine was in general well received. By 1810, Denmark implemented a vaccination program that was mandatory and free of charge. The mandate was enforced at confirmation age (13-15 years), by requiring evidence of immunity from natural smallpox or vaccination for church confirmation. Going forward, ∼ 90% of children were vaccinated, and the mean age at vaccination dropped to ∼1 year of age by the 1820s.^4^ After 1810 smallpox disappeared from Denmark, and physicians wrote that the disease had been defeated. However, in 1824 a smallpox outbreak occurred, after 14 “honeymoon years” with-out the disease. The returning smallpox manifested as a milder disease affecting young adults. The return of smallpox had perhaps been anticipated by the Danish medical establishment, as this phenomenon had been observed in Edinburgh in 1818,^5^ and in England in 1818^6^ and 1819.^7^ There was at the time widespread concern and debate about the reasons for this return of the disease, whether it was even smallpox, and whether the vaccine had failed. The etiology was debated as most cases were adults with a milder “modified” disease, characterized by a smaller number of lesions and low mortality.^8^ The returning smallpox was considered a societal threat and to avoid further transmission in Copenhagen, all cases – mild and severe – were isolated at the Søkvæsthuset hospital in the city.

To investigate the effectiveness of the first smallpox vaccine against infection and death, we searched historical medical journals for studies of post-honeymoon smallpox outbreaks during 1824-1835. From May 1835 onward, the disease was no longer considered a societal threat, and efforts to isolate all cases ended, implying less complete data. We identified five Danish-language publications from the period by the physicians Møhl, Hoppe and Wendt. We also uncovered the patient records from Søkvæsthuset underlying the publications. Analyzing the smallpox data provided by the physicians with modern epidemiological methods we demonstrate that the disease was as deadly as in the pre-vaccination era among the unvaccinated, and that the vaccine was highly protective against mortality.

However, the protection of the single-dose childhood vaccination against smallpox infection waned after a decade or two, explaining the changing age distribution and milder manifestation in the vaccination era.

## METHODS

### Data on smallpox cases and deaths during 1824-1835 outbreaks

Patient records of Søkvæsthuset.^7^ Søkvæsthuset recorded the name, admission date, address, age, and outcome of all patients. Vaccination status was not reported in the surviving version of the list. As both mild and severe smallpox cases were admitted to the hospital for quarantine, we assume these records include most smallpox cases in Copenhagen during 1824-1835. After July 1835, Søkvæsthuset closed because smallpox was no longer considered a societal threat. Afterwards only severe cases were admitted, with patients distributed across several city hospitals.^7^ During 1824-1835, a total of 3795 smallpox patients were admitted. Of these, 186 died (CFR ∼5%). We used these data to compute the age distribution of smallpox cases in the 1824-1835 period (Table 1).

**Table 1.** **Age shift** in smallpox before and after the vaccine introduction. Data from Søkvæsthuset records of annual smallpox cases admitted in Copenhagen.^7^

| ***Epidemic period<br>(year range)*** | *No. adult cases<br>≥ 20 years of age* | *All<br>cases* | *% adult cases<br>≥ 20 years of age* |
|---|---|---|---|
| ***before 1800*** |  |  | *2% of deaths\** |
| ***1824-1826*** | *397* | *1079* | *37%* |
| ***1828-1830*** | *326* | *564* | *58%* |
| ***1833-1835*** | *819* | *1259* | *65%* |

\* Based on tabulated mortality data from the records of Helligaand Parish, Copenhagen, 1730-39 and 1790-1799<sup>18</sup>.

Contemporary publications. We identified five publications that analyzed outbreak data from the epidemics during 1824-1835, and which had data on vaccination status. These include four papers written by the physicians Møhl and Hoppe, who attended the smallpox patients at Søkvæsthuset, and a book by Wendt. These publications include data from each of the five smallpox outbreaks in Copenhagen in 1824,^6^ 1825-1827 ^10^, 1828-30^11^, 1830-32^12^, and 1835^3^. The 1835 outbreak data are from Almindeligt Hospital for July-Dec 1935 and thus were collected after the isolation policy of milder cases was no longer in place.

### Computing Case Fatality (CFR) and Vaccine Effectiveness (VE)

Using summary data from tables of data extracted from these publications for each of the five smallpox outbreaks during 1824-1835, we computed the CFRs by vaccine status and the vaccine effectiveness (VE) against mortality as an outcome. For the 1835 outbreak Wendt only tabulated data from May 15-December when the policy of isolating mild cases was no longer in place. We used summary tables from the five publications by Møhl, Hoppe, and Wendt to compute CFRs for patients of different ages.

CFR for vaccinated:CFR^vax^=deaths^vax^/cases^vax^

CFR for unvaccinated: CFR^unvax^=deaths^unvax^/cases^unvax^Vaccine effectiveness: VE =1-(CFR^vax^/CFR^unvax^), with 95% confidence intervals.

Due to the VEs being relations between fractions where the fractions sometimes involve small numbers, the relative risks and confidence intervals were computed using Koopman’s method.^10^

## RESULTS

### Analysis of data from five smallpox outbreaks in Copenhagen during 1824-1835

After the introduction of the mandatory vaccination program in 1810 a “honeymoon period” free of smallpox followed. But then the disease returned to the city in December 1823 and caused five smaller, less deadly outbreaks between 1824-1835.^1^ These were documented by the physicians Møhl, Hoppe and Wendt in five publications.^8,10–12,14^

The 14-year-long “honeymoon period” ended when in December 1823, a 30-year-old worker who may have encountered smallpox on a ship in the Copenhagen harbor was hospitalized at Almindeligt Hospital for the care of the sick and poor. A hospital-based out-break followed, as curious medical students wanting to see the rare illness fell ill, along with other patients at the hospital. Soon smallpox reached a poor house (an institution for paupers) and spread all over the city. To avoid further spreading all patients diagnosed with smallpox were admitted to a dedicated smallpox hospital, Søkvæsthuset. The physician Møhl was ordered to be quarantined at Søkvæsthuset in 1824 where he cared for the patients throughout this and the subsequent outbreak in 1825-1827. In 1825 he published an article about the 1824 outbreak where he had treated and studied all mild and severe smallpox cases in the city. He described the new and milder clinical presentation of “modified” smallpox and argued that it could be explained by waning vaccine protection ^6^. In particular, he noted that very few vaccinated children below the age of six got smallpox, whereas smallpox infections became more common in older cohorts. He also noted that none of the patients were older than 40 years and suggested that this was due to lifelong protection from surviving smallpox in their childhood in the late 1700s. From this, he deduced that vaccine-induced immunity waned over time:

*“From this table [of numbers of cases] one sees how much larger the number of adults, who were vaccinated many years ago, was in comparison to the number of children. (*…*) one can hardly refrain from drawing from these facts the following conclusion: that the protective power of the vaccine is diminished over the years (*…*)” - Møhl 1825*^8^, *p. 194 (our translation from Danish)*

The second outbreak lasted from 1825-1827 and peaked in May 1826. Møhl admitted 616 patients whereof 35 died. Smallpox returned in December 1827, first in poor houses and in a prison population. This epidemic lasted throughout 1828-1830 and peaked in Sept 1829. He admitted 562 patients to the hospital and 29 of them died.^11^ In 1830, Dr Møhl died for unrelated reasons and was replaced by Dr Hoppe who reported on the data Møhl had collected during the following 1828-1830 epidemic.^11^ Later Hoppe reported on a fourth out-break in 1832-1834.^11^ A fifth outbreak occurred in 1835 and peaked in Nov that year.^3^ For this outbreak we do not have the total number of smallpox cases but city statistics show that 434 persons died of the disease.^3^ The only surviving data we could find were from Almindeligt Hospital and represented the 2^nd^ half of the 1835 epidemic after Kvæsthuset had closed.^3^ As the mandatory isolation of patients with mild infections had ended by May 1835, the CFRs are here greater and less reliable than in previous outbreaks because some proportion of mild cases are not included. The data we analyzed from Almindeligt Hospital included 99 deaths (around 25% of all smallpox deaths in the city). We have included these data because this was the only outbreak for which double-stratified data by age and vaccination status was available to us, so that we could compute the CFR trends over time since childhood.

### Case Fatality (CFR) and Vaccine effectiveness (VE)

For each of the five smallpox outbreaks during 1824-1835, we computed the CFR for vaccinated and unvaccinated cases using tabulated data in the five publications by Møhl, Hoppe and Wendt (Table 2). For example, from the 1824 outbreak data we could extract the numbers of vaccinated and unvaccinated persons, broken down by outcome (died or lived). From this we computed the CFR in the vaccinated patient group to be 0·4% (1/248). For the unvaccinated cases the CFR was 25·7% (39/152). From these CFR estimates we computed the relative risk (RR) of dying among all smallpox cases and calculated the vaccine effectiveness against death (VE) to be 98%. We then repeated this calculation for each outbreak and found the same pattern: high CFR in unvaccinated patients and a high VE.

**Table 2.**
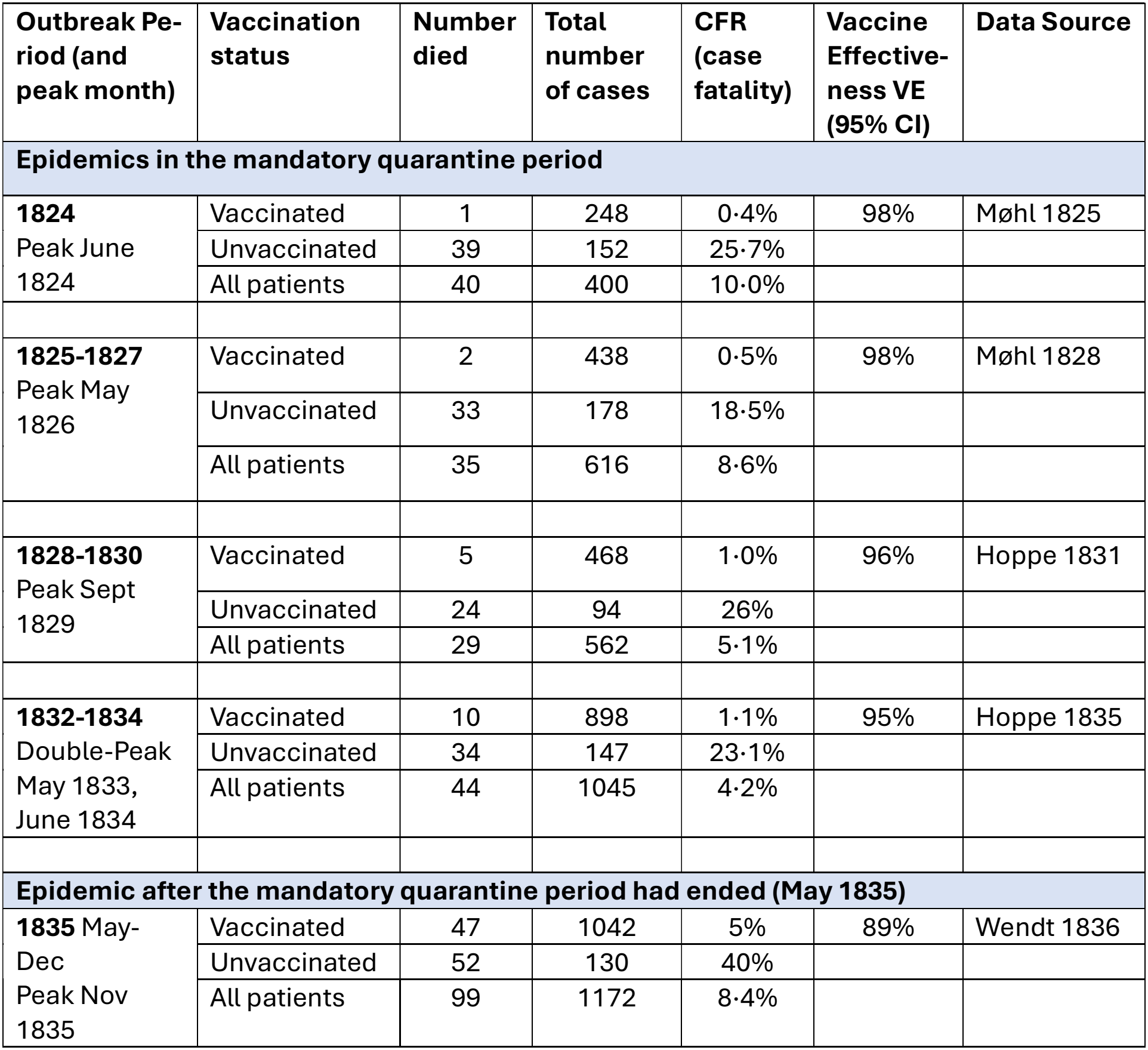
**Data from 5 smallpox outbreaks in Copenhagen** during 1824-1835 presented in five publications by physicians Møhl, Hoppe and Wendt.^1,6,8,9,11^ From numbers of cases and deaths we computed the Case Fatality (CFR) and vaccine effectiveness. The peak month of the outbreak is indicated.

| **Outbreak Pe-riod (and peak month)** | **Vaccination status** | **Number died** | **Total number of cases** | **CFR (case fatality)** | **Vaccine Effective-ness VE (95% CI)** | **Data Source** |
|---|---|---|---|---|---|---|
| **Epidemics in the mandatory quarantine period** |  |  |  |  |  |  |
| **1824**<br>Peak June 1824 | Vaccinated | 1 | 248 | 0·4% | 98% | Møhl 1825 |
|  | Unvaccinated | 39 | 152 | 25·7% |  |  |
|  | All patients | 40 | 400 | 10·0% |  |  |
| **1825-1827**<br>Peak May 1826 | Vaccinated | 2 | 438 | 0·5% | 98% | Møhl 1828 |
|  | Unvaccinated | 33 | 178 | 18·5% |  |  |
|  | All patients | 35 | 616 | 8·6% |  |  |
| **1828-1830**<br>Peak Sept 1829 | Vaccinated | 5 | 468 | 1·0% | 96% | Hoppe 1831 |
|  | Unvaccinated | 24 | 94 | 26% |  |  |
|  | All patients | 29 | 562 | 5·1% |  |  |
| **1832-1834**<br>Double-Peak May 1833, June 1834 | Vaccinated | 10 | 898 | 1·1% | 95% | Hoppe 1835 |
|  | Unvaccinated | 34 | 147 | 23·1% |  |  |
|  | All patients | 44 | 1045 | 4·2% |  |  |
| **Epidemic after the mandatory quarantine period had ended (May 1835)** |  |  |  |  |  |  |
| **1835** May-Dec<br>Peak Nov 1835 | Vaccinated | 47 | 1042 | 5% | 89% | Wendt 1836 |
|  | Unvaccinated | 52 | 130 | 40% |  |  |
|  | All patients | 99 | 1172 | 8·4% |  |  |

For the first four outbreaks we found profoundly different mortality risk in the vaccinated and unvaccinated. The CFR of the unvaccinated was ∼25%, comparable to the high severity of smallpox in the pre-vaccination era, while CFR ∼1% for the vaccinated. For the 1835 outbreak (where we only have data from July to December after the mandatory quarantine was lifted and only severe cases were hospitalized), the CFRs were somewhat higher.

The vaccine effectiveness remained high during 1824-1835, but declined gradually, from 98% to 89% over the 11-year study period. There was also a clear tendency for the number of cases in the vaccinated to increase with age (Table 1). This pattern of waning vaccine protection was not lost on Møhl and Hoppe, as shown by the above quote.

### Age distribution of smallpox cases

The data in Table 1 show the age shift of smallpox from a childhood illness to a disease of adulthood. In the 1824 outbreak, 37% of the cases were in adults (Table 1). When stratifying by vaccination status, one sees an increasing number of adult vaccinated smallpox cases from age one to 23 years and none over 23 years of age. This is as expected, given that the earliest vaccinated cohort of children were born in 1801 and thus were 23 years old in 1824 (Figure 1). The increasing number of vaccinated cases with age is consistent with waning immunity in those who had been vaccinated long ago. Møhl commented already in his 1826 and 1828 works that those who had been vaccinated 6-20 years earlier could become infected, albeit with modified/milder form of illness. In contrast, the age distributions of unvaccinated smallpox cases were bimodal and highest among young children, likely because not all young children had been vaccinated by the age of two (Figure 1). A second peak of cases in young adults around 24 years of age is best explained by a “lost generation” born around 1800 -- too early to have benefitted from the vaccination program and too late to have protection from surviving smallpox infection in childhood as the disease came under vaccine control (Figure 1).

**Figure 1.**
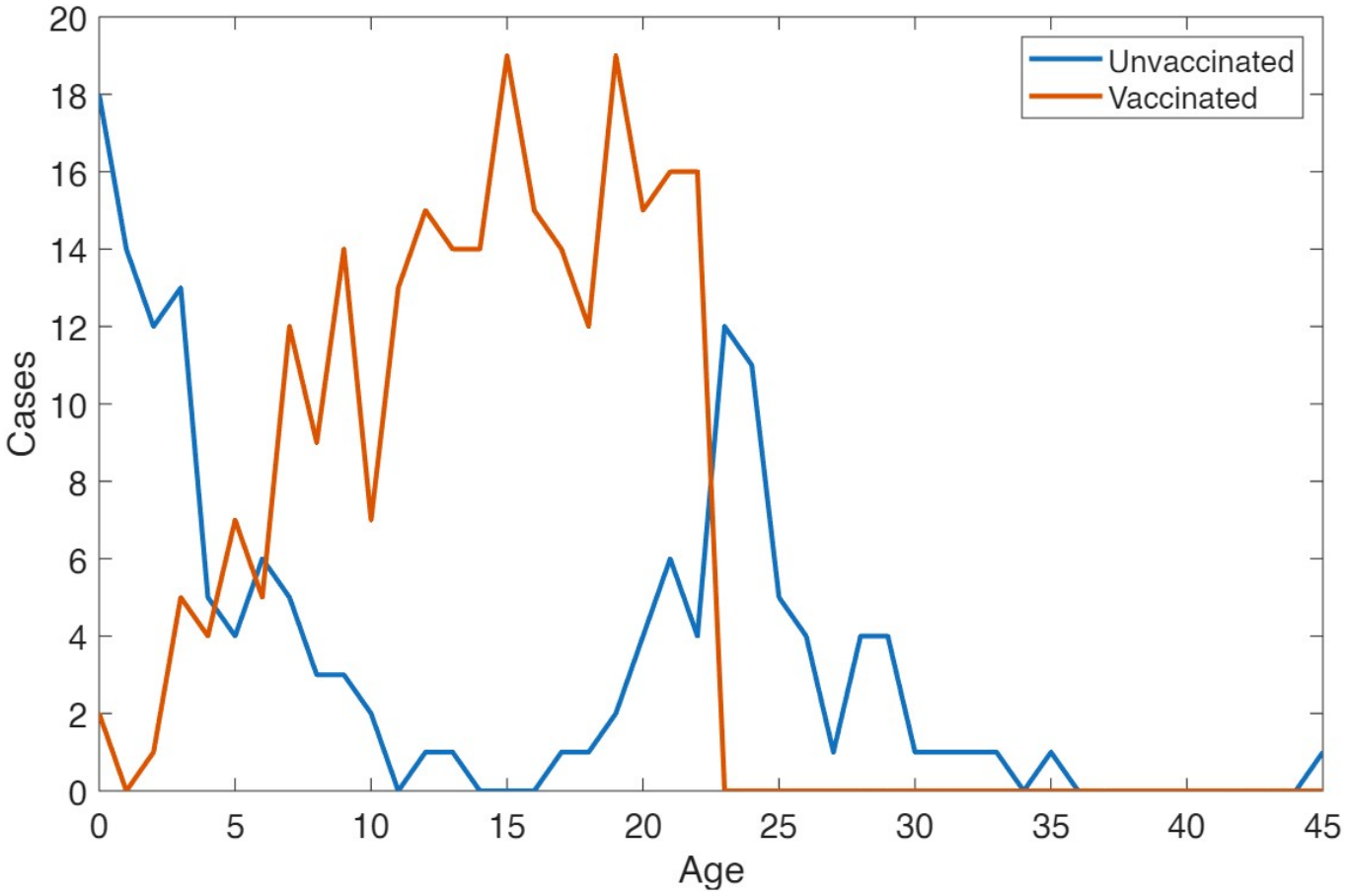
**Age distribution** of smallpox cases in the 1824 epidemic, by vaccination status. The increase in vaccinated cases with age shows vaccine waning. Data obtained from Møhl.^6^

### Age variation in CFR and VE

Data from the 1835 outbreak^3^ can be used to study the waning of vaccine protection over time. In Almindeligt Hospital, 98 smallpox patients died (47 vaccinated and 38 unvaccinated) out of 1165 admitted smallpox cases. This patient cohort accounted for ∼25% of all smallpox deaths in Copenhagen during 1835,^3^ and within the cohort ∼90% of persons born after 1810 were vaccinated.^7^

From a data table from Wendt ^3^ detailing data from 1042 cases with age, vaccine and out-come, we computed the case fatality and vaccine effectiveness (VE) for each ten-year age group. Across all ages, the CFR was 4·5% among the vaccinated and 41% among the unvaccinated (Table 3). Among the unvaccinated, the CFR was similar in all age groups (from 38% in the 40-49-year-olds up to 54% in those under 10). For the children less than 10 years old, the relative risk was not defined for the vaccinated group (zero deaths). For the unvaccinated, the CFR of those under 10 years old had very wide confidence intervals. Therefore, we combined the age groups <20 years and estimated a VE of 95%. The VE was significantly lower for the 30-39 age group, demonstrating vaccine waning with age (time since childhood vaccination). The 40-49 age group had too few cases to obtain significant CFR and VE estimates. Across all the outbreaks, very few cases were older than 45 years. This we interpret as those born before 1800 being protected for life by immunity from natural infection in childhood. The 1835 CFRs are necessarily overestimated as many milder cases likely went unnoticed due to the abolition of mandatory isolation. Un-derestimation of the number of cases likely affected CFR estimates for the vaccinated more than the unvaccinated due to the typically milder cases in the vaccinated population. This would have resulted in an underestimation of VE. However, this cannot explain the rising CFRs with age among the vaccinated, nor the fall in VE with age (Table 3).

**Table 3.** **Effect of vaccination and waning** in decades since vaccination. Age-specific Case Fatality (CFR) and Vaccine Effectiveness (VE) for 10 year age groups among smallpox cases admitted to Almindeligt Hospital in Copenhagen during May-Dec 1835 (after quarantine policy was lifted). *The 40-49 year age group results are difficult to interpret due to small numbers of cases.

|  | **Vaccinated** |  |  | **Unvaccinated** |  |  | **Vaccine effectiveness** |  |
|---|---|---|---|---|---|---|---|---|
| **Age range (years)** | Cases | Deaths | **CFR_vax** | Cases | Deaths | **CFR_unvax** | **VE** | 95%CI |
| **<10** | 11 | 0 | **0%** | 13 | 7 | **54%** | **100%** | 49-100% |
| **10-19** | 244 | 5 | **2%** | 17 | 7 | **41%** | **95%** | 83-99% |
| **20-29** | 640 | 21 | **3%** | 13 | 6 | **46%** | **93%** | 85-97% |
| **30-39** | 144 | 19 | **13%** | 72 | 30 | **42%** | **68%** | 48-81% |
| **40-49\*** | 3 | 2 | **67%** | 21 | 8 | **38%** | **-75%** | -287-51% |
| **All Ages** | 1042 | 47 | **5%** | 123 | 51 | **41%** | **89%** | 87-92% |

## DISCUSSION

Using unique historical reports by Danish physicians from smallpox outbreaks occurring during 1824-1835,^1,6,8,9,11^ we investigated how the vaccine affected the CFR and age distribution of smallpox cases in Copenhagen when the disease returned after a 14-year honeymoon period between 1810 and 1824. We find that the vaccine almost totally prevented mortality (98% effectiveness against death in 1824, declining to 89% by 1835). Among the vaccinated, the risk of being infected increased with age, consistent with vaccine waning. In a previous paper, we likewise used a dynamic model to show that vaccine waning may explain the age shift of smallpox after 1824 ^4^. Here we confirm this conclusion, this time based on direct study of vaccinated individuals and their time since vaccination. While after 1824 the disease was milder in the large majority of individuals who were vaccinated, the data showing a CFR of 25% among the unvaccinated cases clearly demonstrate that the virus was still as virulent as before 1810.

From our data from historical Copenhagen it is evident that in a well vaccinated population where the vast majority of cases are breakthrough infections in the vaccinated, even a highly virulent variola virus (later termed *Variola major*) would cause a mild outbreak, making it appear that the pathogen itself had become less virulent. Indeed, the physicians at the time entertained the possibility that the returning smallpox was in fact a new, inherently milder disease. While we have documented that this was not the case, starting in the early 20th century, physicians around the world noted that some outbreaks of smallpox had much lower CFR than others, and the responsible smallpox strain was eventually termed *Variola minor* (or Alastrim). Our work shows that it is impossible to distinguish outbreaks of inherently low virulence (V. minor) and V. major outbreaks unless the vaccination coverage in the population is taken into consideration. For some reason this is rarely the case in literature. While it is in some cases indisputable that less severe variants did exist,^15,16^ out-breaks were usually classified as one or the other based solely on population level CFR.^17^

Our study follows the comprehensive analysis of smallpox in Sweden by Peter Sköld.^1^ He also found that the vaccine was associated with a sharp reduction in smallpox mortality after the introduction of the vaccine, and that, after a “honeymoon period” without outbreaks, smallpox returned as a less deadly disease of adulthood.^6^ Whereas the seminal study by Sköld was based only on mortality data, the Danish data from 1824-1835 are at record-level and contain information about all cases, age, vaccination status and outcome (death). This, and the contemporaneous treatment of smallpox as a societal threat so that all cases were isolated at a single hospital, allowed us to study the CFR of smallpox when it returned after the honeymoon period. Our Danish data confirm and strengthen his findings on the effect of the vaccine on smallpox epidemiology. In the present case, two thirds of smallpox patients were adults aged 20 or more by 1833-35, an even higher proportion than the one found in Sköld’s mortality data.

Our present results likewise echo the observations made already in 1825 by Dr Møhl who noted the disappointment over the return of smallpox after the honeymoon period. Despite this, he described how the severity of smallpox was by then far less than before the vaccine was introduced. He proceeded to conclude that the vaccine protected against severe illness and mortality with protection waning after 6-20 years against illness and later against mortality.^9,11^ He referred to British colleagues who had discussed a similar return of smallpox in London two years earlier,^11^ noting much contemporary debate about the etiology of “modified smallpox”, and even suggestions that this was a new disease. However, the strong link made by Møhl between being vaccinated and getting a “modified” course of smallpox made him conclude that the etiology of modified and natural smallpox was the same. Using modern statistical methods we here confirm his observation.

Our high CFR estimate for the unvaccinated is in apparent contradiction with Schneider and Davenport (2026)^2^ who recently used smallpox mortality data from Sweden to conclude that the CFR of smallpox before vaccination was 8-10%.^2^ As they estimate this by a comparison of population-wide smallpox mortality with the age-specific infection rates required to induce high immunity in adults, this estimate is in our opinion really an *infection* fatality rate (IFR). We agree that to determine how deadly the disease was in the general population before the vaccine this is the right approach to take, and we will conduct a similar study using the Danish data in a forthcoming paper.

In summary, we found that the vaccine was highly protective, but that this protection waned over time so that persons could again become infected with smallpox after a decade or so. The vaccine-induced protection, especially against infection but also increasingly against death, was significantly weakened in those vaccinated over 20 years earlier. Our CFR analysis shows conclusively that unvaccinated persons who had not had natural smallpox in their childhood continued to suffer severe smallpox, similar in severity to the epidemics before the vaccine era. This shows that the virus itself had not changed after 1800. In addition to the drop in CFR, smallpox also changed from a disease of childhood to adulthood after the 1810-24 honeymoon period. We explain this age shift by two factors: firstly, the “lost generation” of young adults born before 1800 as described in the introduction. Secondly, the contribution of the waning of vaccine protection over time. We have previously used a mathematical model to explain the temporal epidemic dynamics and age patterns due to vaccine waning,^4^ and here we repeat this result using statistical data analysis alone.

In our present study, we have shown that the introduction of the smallpox vaccine in Copenhagen in 1801 caused the age distribution of smallpox cases to shift and reduced its apparent severity, to the point where it was possible for contemporaries to suggest that it was no longer the same illness. However, we also show that smallpox returned in 1824 due to waning vaccine immunity and a susceptible “lost generation”. The currently observed resurgence of some infectious diseases due to limited vaccine duration and unvaccinated segments of the population thus have a precedent dating back to the earliest vaccination program. More optimistically, the potential massive benefits of vaccination programs for public health were likewise already apparent at the time. As Hoppe said,

> *“It is in no way impossible that we, by widespread and early immunization (…) could become – not free of smallpox, this is something we never should expect as susceptibility returns too early – but free from deaths due to smallpox*.*” - Hoppe, 1831*

Hoppe and his colleagues would be pleased to know that smallpox was eventually eradicated by a massive, global WHO vaccination effort in 1979. This final victory occurred about a century after the mandatory vaccination program had ended the disease in Denmark.

## Author contribution statement

AE contributed to the analysis, produced the figure, and co-wrote the manuscript. SP located and transcribed historical sources and provided the necessary information on the historical context. MJFG contributed biological and genetic information and managed references. JBP provided further historical information on the disease history of smallpox before the period of this study. AGP likewise contributed genetic and phylogenetic information. RD contributed methods and background information on estimating smallpox CFRs from historical sources. LS initiated the project, located relevant papers, carried out the mathematical and statistical analyses, produced the tables, and co-wrote the manuscript. All authors edited and approved the manuscript.

## Data availability statement

All data used here are publicly available in the cited sources.^1,6,8,9,11^ Furthermore, the transcribed patient records from Søkvæsthuset can be found on GitHub (<https://github.com/basspoder/smallpox_post_honeymoon_phase_1824_1872/releases/tag/Dataset>).

